# Profiling COVID-19 pneumonia progressing into the cytokine storm syndrome: results from a single Italian Centre study on tocilizumab versus standard of care

**DOI:** 10.1101/2020.05.01.20078360

**Authors:** Luca Quartuccio, Arianna Sonaglia, Dennis McGonagle, Martina Fabris, Maddalena Peghin, Davide Pecori, Amato De Monte, Tiziana Bove, Francesco Curcio, Flavio Bassi, Salvatore De Vita, Carlo Tascini

## Abstract

**Objective:** Approximately 5% of patients with coronavirus disease 2019 (COVID-19) develop a life-threatening pneumonia that often occurs in the setting of increased inflammation or “cytokine storm”. Anti-cytokine treatments are being evaluated but optimal patient selection remains unclear, and the aim of our study is to address this point.

**Methods:** Between February 29 to April 6, 2020, 111 consecutive hospitalized patients with COVID-19 pneumonia were evaluated in a single centre retrospective study. Patients were divided in two groups: 42 severe cases (TOCI) with adverse prognostic features including raised CRP and IL-6 levels, who underwent anti-cytokine treatments, mostly tocilizumab, and 69 standard of care patients (SOC).

**Results:** In the TOCI group, all received anti-viral therapy and 40% also received glucocorticoids. In TOCI, 62% of cases were ventilated and there were 3 deaths (17.8±10.6 days, mean follow up) with 7/26 cases remaining on ventilators, without improvement, and 17/26 developed bacterial superinfection. One fatality occurred in the 15 TOCI cases treated on noninvasive ventilation and 1 serious bacterial superinfection. Of the 69 cases in SOC, there was no fatalities and no bacterial complications. The TOCI group had higher baseline CRP and IL-6 elevations (p<0.0001 for both) and higher neutrophils and lower lymphocyte levels (p= 0.04 and p=0.001, respectively) with the TOCI ventilated patients having higher markers than non-ventilated TOCI patients.

**Conclusion:** Higher inflammatory markers, more infections and worse outcomes characterized ventilated TOCI cases compared to ward based TOCI. Despite the confounding factors, this suggests that therapy time in anti-cytokine randomized trials will be key.

**Funding:** This research received no external funding.

**Conflicts of Interest:** “The authors declare no conflict of interest.”

**Highlights:**

- There is an urgent need for markers of prognosis in COVID-19.
- Higher inflammatory markers best select tocilizumab treatment.
- The ward based tocilizumab group showed better responses and less infections than ICU tocilizumab group.
- The former group may be the best for evaluating the impact of anti-cytokine therapy in COVID-19.
- The known poor risk factors for COVID-19 infection were present in the TOCI treated rather than in the good prognosis standard of care group.

## Introduction

The outbreak of novel coronavirus 2019 (COVID-19) caused by severe acute respiratory syndrome coronavirus 2 (SARS-CoV-2) is a global pandemic [1]. About twenty-five percent of patients have a seriously ill disease. A fraction of them may develop a very severe pneumonia which may progress to acute respiratory distress syndrome (ARDS) or end-organ failure that may be associated with a cytokine storm syndrome [2]. Laboratory features associated with ARDS or death included neutrophilia, coagulation dysfunction [e.g., higher lactate dehydrogenase (LDH) and D-dimer] [3]. Markedly high levels of interleukin (IL)-2R, IL-6, IL-10, and TNF-α and the absolute numbers of CD4+ and CD8+ T lymphocytes being markedly low seem to characterize the most severe cases [4]. Starting from the first preliminary experience on the apparent efficacy of tocilizumab in COVID-19 pneumonia [5], many multicenter trials are ongoing to test anti-cytokine treatments in critically ill patients.

Nevertheless, robust data to predict the outcome of COVID-19 pneumonia after the hospital admission are still lacking [6], though they are urgently needed in order to facilitate the assessment of anti-cytokine treatment efficacy in worse prognosis patient groups and not milder disease. The aim of this retrospective study was to evaluate baseline laboratory and immunological features in patients hospitalized for COVID-19 pneumonia and to explore such parameters in relationship to standard of care (SOC group) therapy versus anti-cytokine therapy, mainly tocilizumab, (TOCI group) that was mostly used either in ventilated patients in the ICU or non-invasively ventilated patients, mostly in the ward setting. Our single centre experience and approach showed that the milder hospitalized SOC group faired well as did cases with cytokine storm treated with tocilizumab outside of the ICU setting without ventilator support. Severe complications including bacterial infections complicated tocilizumab in the ICU setting but not ward-based tocilizumab therapy. Therefore, randomized trials should target non-ICU patients to prevent cytokine storm evolution.

## Methods

This study was undertaken to identify laboratory features for more serious COVID-19 disease (i.e., to determine which cases that might theoretically benefit from anti-cytokine drugs). In this monocentric retrospective case-control study, the clinical and immunological characteristics of 111 consecutive patients with COVID-19 were analyzed. Patients were admitted to our hospital from February 29 to April 6, 2020. All but 6 patients presented to our hospital with 6 cases transferred from three other hospitals (all of whom eventually received tocilizumab).

Oral or written consent was obtained from patients. The study was conducted in accordance with the ethical principles of the Helsinki Declaration and ethical approval was given by local Ethics Committee (CEUR-2020-Os-102).

Besides clinical evaluation, the level of CRP and IL-6, when available, guided the decision towards anti-cytokine treatments. Clinical decisions for the treatment of all these patients were taken usually within the first week after the admission, and during this time, the laboratory tests were repeated. Demographic, clinical and laboratory characteristics, treatments and outcome data were collected. Identification of cases of COVID-19 virus was based on the detection of unique sequences of virus RNA by nucleic acid amplification tests (NAAT) such as RT-PCR with confirmation by nucleic acid sequencing. The following genes were investigated: E gene for screening and then RdRp and N genes of SARS-CoV-2 for confirmation [7].

Some laboratory data analysed at the admission are reported in table 1, including flow cytometry analysis with antibodies for the following subpopulations: CD19+ B cells, CD3+CD4+ T cells, CD3+CD8+ T cells, CD56+ NK cells, platelet count (cell/microL) and serum IL-6 (pg/ml), measured by CE_IVD electrochemiluminescence immunoassay (Elecsys IL6, Cobas, physiological range < 7pg/ml) with results being available within 48 hours.

Variables were reported as mean and standard deviation or median and interquartile range (IQR), as appropriate, or frequency rates and percentages if categorical; consequently, comparisons between TOCI and SOC groups were made by parametric tests (t-test for two independent samples) or no parametric tests (Mann-Whitney test) for continuous variables. Proportions were compared by χ2 test, or Fisher exact test. Bivariate correlation was made by two tailed Pearson or Spearman tests. All statistical analyses were performed using SPSS version 15.0 software (SPSS Inc). For unadjusted comparisons, a 2-sided α of less than .05 was considered statistically significant. No corrections were made for multiple comparisons due to the explorative nature of the study.

When the laboratory parameters were available, the patients were classified into two groups: the first group comprised 42 cases who developed a serious COVID-19 disease that were deemed suitable for tocilizumab 8 mg/kg intravenously as a single infusion. In TOCI failures, two patients were then treated with anakinra 200 mg/day subcutaneously for three consecutive days. A second group of 69 cases who received supportive therapy [standard of care group (SOC)] comprised those initially admitted to the hospital for COVID-19, and who were treated with SOC based on clinical and laboratory features (Table 1).

**Table 1.**
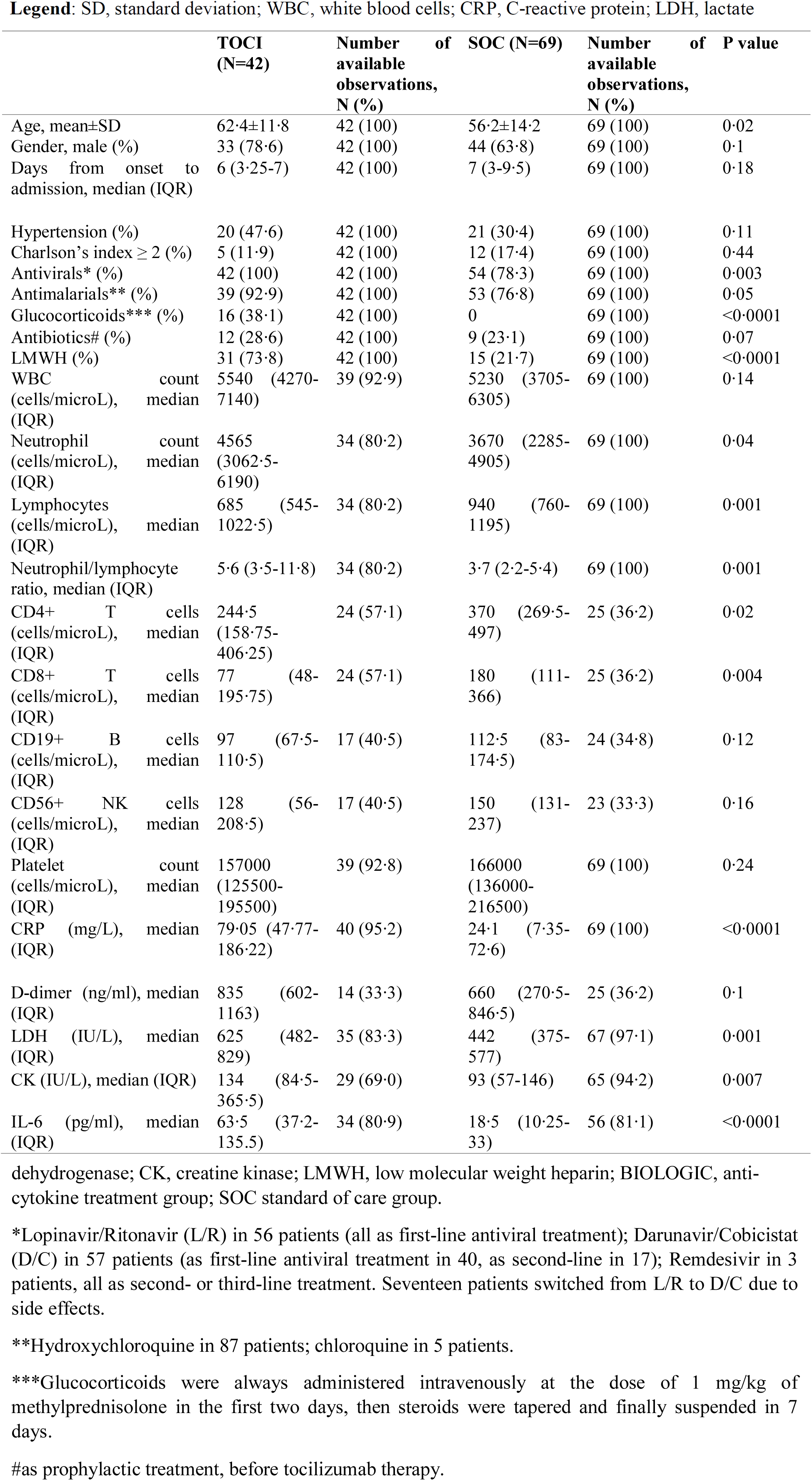
Main comparisons between treatment groups at day 0 (hospital admission).

## Results

### Patients’ characteristics and outcome

Table 1 reports the main demographic and clinical features of the two groups. Patients were predominantly male (77/111, 69.4%) with a mean age of 58.5±13.6 years. Patients in TOCI were slightly older than SOC (p=0.02) (table 1). Globally, at the hospital admission, resting oxygen saturation equal or below 93% was available for 45 patients (40.5%).

Antiviral treatments were employed in 100% of TOCI group and 80% of SOC group (Table 1). Notably, nearly 40% of TOCI group received glucocorticoids but none of the SOC group did (Table 1). There was no difference between groups regarding the time of reaching a negative swab test (supplemental file).

Among TOCI group, 18 (43%) patients were originally referred to the Infectious Disease Unit with 3 being subsequently transferred to ICU before tocilizumab administration (Figure 1) with 24/42 patients (57%) ICU transfers within 24 hours of hospital admission. The majority of patients received tocilizumab in the ICU (27/42, 64.3%) with the remaining 15 cases receiving TOCI on the ward. Tocilizumab was administered after a mean time of 8.4±3.7 days from disease onset as addon treatment. Of the 27 patients that were transferred to ICU, 26 (96.3%) were intubated with subsequent tracheostomies in 8 (7.2%), while only one was on noninvasive ventilation.

**Figure 1.**
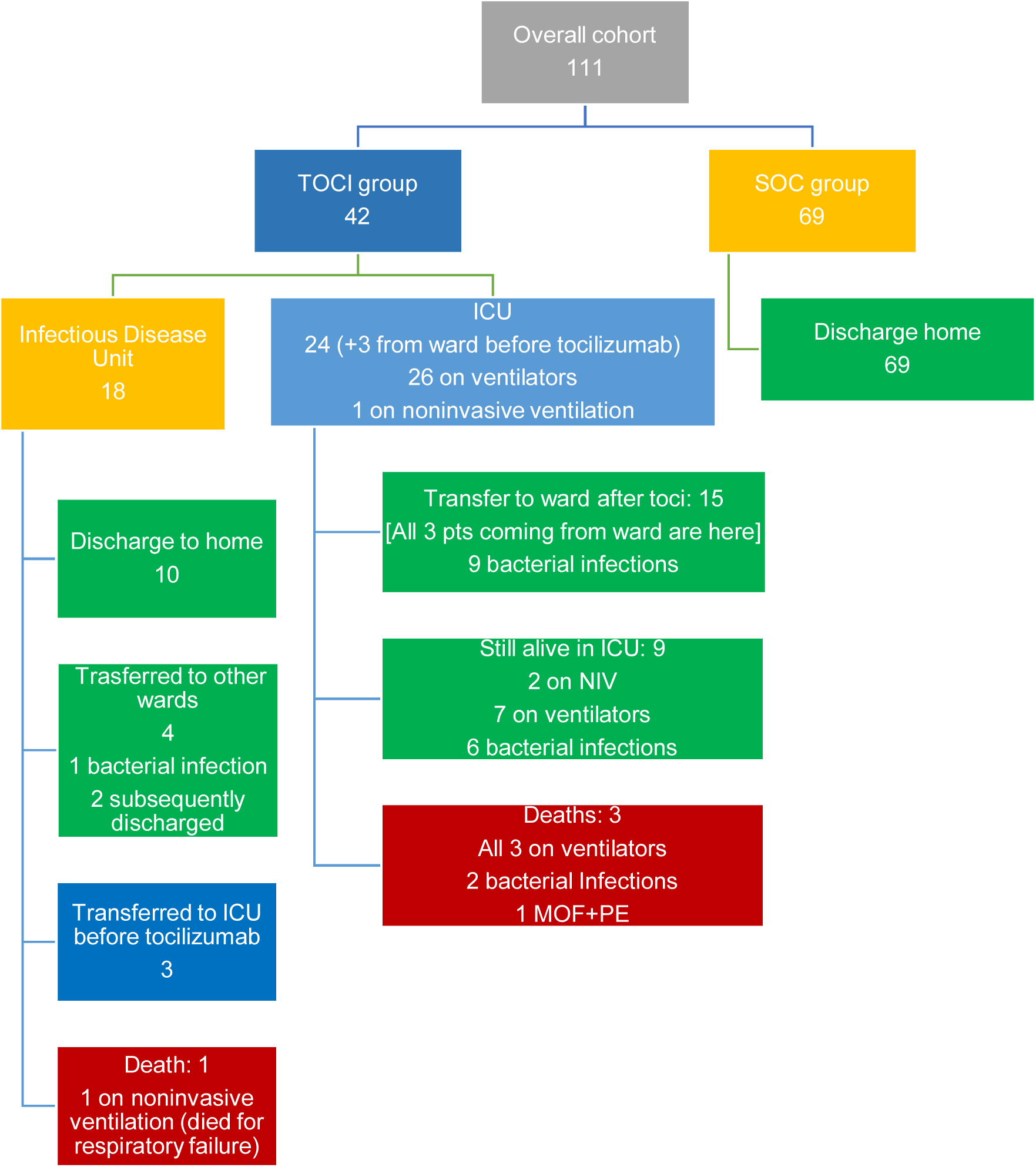
The chart illustrates the outcomes of the two treatment groups. **Legend:** TOCI, anti-cytokine group; SOF, standard of care group; MOF, multi-469 organ failure; ICU, 470 intensive care unit; NIV, noninvasive ventilation; PE, pulmonary embolism.

There were no fatalities in the SOC group (Figure 1). Overall, at April 18, 2020, 4/42 TOCI patients had died (9.5%). Of the TOCI ventilated patients 15/26 (57.7%) had a good outcome. When combined with fatality rate, 11/26 (42.3%) patients in the TOCI ventilated group can be deemed as non-responders. By contrast, 15/16 (93.7%) TOCI non-ventilated patients can be deemed as responders with a single fatality (Figure 1). Importantly, at the hospital admission, TOCI patients who required invasive ventilation showed higher levels of inflammation markers, higher LDH and lower lymphocyte count than non-ventilated TOCI patients (Table 2).

**Table 2.**
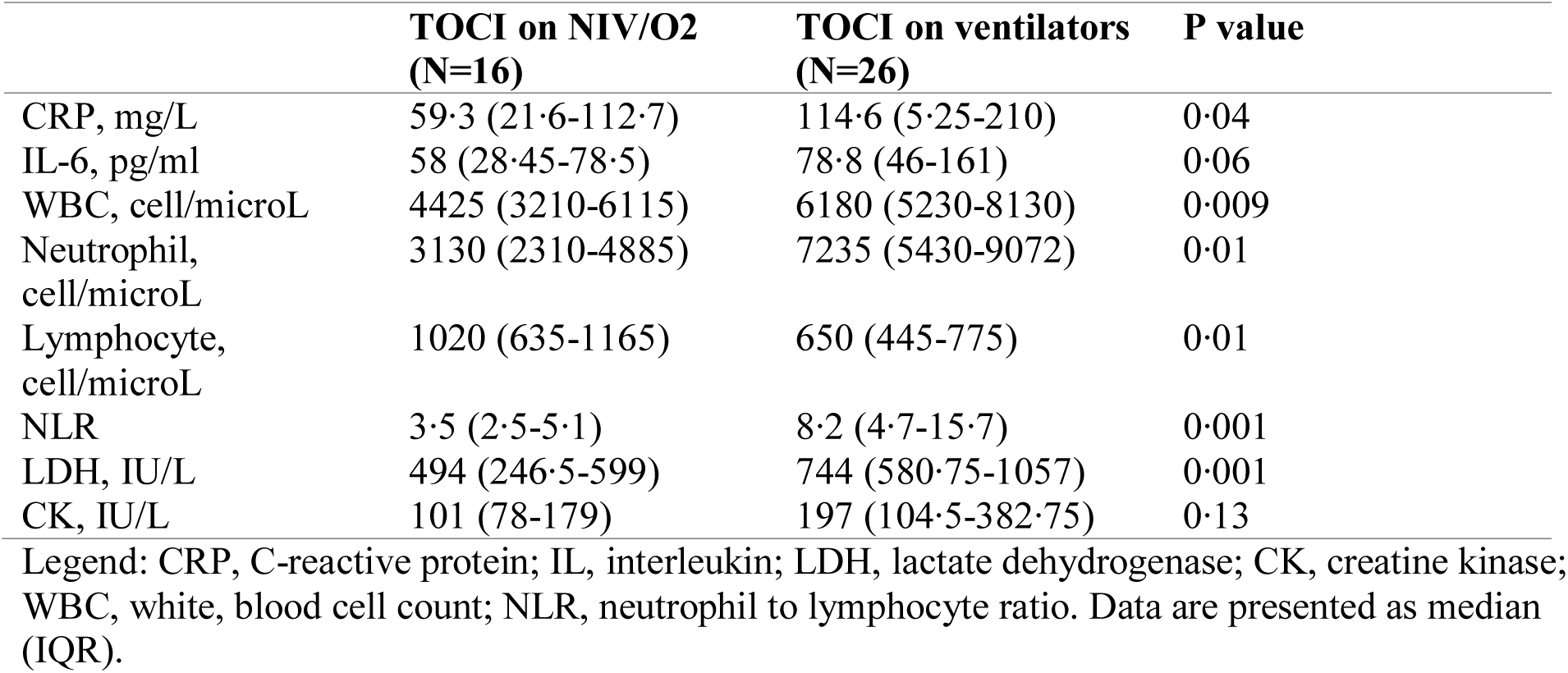
Laboratory marker comparison between the two Tocilizumab treated subgroups.

Eighteen out 111 patients (16.2%) experienced bacterial superinfection that were almost exclusively in the TOCI group (Figure 1). Three out of four deaths and 17/18 bacterial complications occurred in ICU (all 3 deaths as well as all the bacterial complications occurred in patients on ventilators or in the non-ventilated TOCI group (Figure 1).

While all the patients in the SOC group recovered, in the TOCI group, 9/42 (21.4%) patients completely recovered, and 21/42 (50%) patients showed a clear and rapid improvement after tocilizumab. A rapid improvement on anakinra after tocilizumab occurred in one case. In the 21 recovered TOCI treated group complicating infections arose in 11 (52.4%). In the remaining 12 non-responder patients, four of them died, including one treated with anakinra after tocilizumab failure, and almost all showed co-morbidities including hypertension, obesity, ischemic heart disease or diabetes, or experienced superinfections, which substantially complicated the course.

### Retrospective laboratory marker comparison between treatment groups

At hospital admission, TOCI group showed a significantly higher level of systemic inflammation as resulted by the significant difference of CRP levels [mg/L, median (IQR)] [79.05 (47.8-186.22) vs 24.1 (7.3-72.6) p<0.0001)], and IL-6 levels [pg/mL, median (IQR)] [63.5 (37.25-135.5) vs 18.5 (10.25-33), p<0.0001]. Also, some other laboratory features mirrored a higher level of systemic disease and organ damage in TOCI group, such as LDH [IU/L, median (IQR)] [625 (482-829) vs 442 (375-577), p=0.001] and CK [IU/L, median (IQR)] [134 (84.5-365.5) vs 93 (57-146), p=0.007].

The TOCI group showed a significantly higher neutrophil count (cells/microL) [4565 (3062.5-6190) vs 3670 (2285-4905), p=0.04], lower lymphocyte count [cell/microL, median (IQR)] [685 (545-1022.5) vs 940 (760-1195), p=0.001], CD4+ T cell [244.5 (158.75-406.25) vs 370 (269.5-497), p=0.02], CD8+ T cell subpopulation [77 (48-195.75) vs 180 (111-366), p=0.004]. Also, neutrophil to lymphocyte ratio (NLR) was significantly higher in TOCI group than in SOC group [5.6 (3.5-11.8) vs 3.6 (2.2-5.4), p=0.001]. The TOCI group also showed basal higher levels of LDH (p=0.001) and CK (p=0.007), possibly indicating cardiac injury that is a known bad prognostic sign.

Table 3A reports the correlations between CRP levels and the levels of the other biomarkers in the whole population (TOCI+SOC) and in the whole population after excluding those patients with the worst clinical presentation at the admission (N=24). A moderate to high correlation (>0.5) was found between CRP and the following variables: D-dimer, LDH, neutrophil count and NLR (table 2).

By excluding those patients admitted to the ICU within 24 hours (i.e., the most serious) (table 3B), CRP and IL-6 remained statistically significant as discriminant variables between the two groups (Table 3). Yet, correlations between CRP and IL-6, total white blood cell count, neutrophil count, NLR, LDH were still significant (table 3B).

**Table 3A.** Bivariate correlations between CRP levels and other biomarkers.

| Variable | Spearman's rho<br>correlation coefficient | P value | Number of<br>observations, N<br>(%) |
| --- | --- | --- | --- |
| IL-6 | 0.46 | <0.0001 | 90 (81.1) |
| D-dimer | 0.63 | <0.0001 | 39 (35.1) |
| LDH | 0.62 | <0.0001 | 102 (91.9) |
| CK | 0.23 | 0.03 | 94 (84.7) |
| Total WBC count | 0.49 | <0.0001 | 108 (97.3) |
| Neutrophil count | 0.60 | <0.0001 | 103 (92.8) |
| Lymphocyte count | -0.26 | 0.002 | 103 (92.8) |
| NLR | 0.57 | <0.0001 | 103 (92.8) |

Legend: IL, interleukin; LDH, lactate dehydrogenase; CK, creatine kinase; WBC, white blood cell count; NLR, neutrophil to lymphocyte ratio.

**Table 3B.** Bivariate correlations between CRP levels and other biomarkers by excluding those patients transferred to ICU within 24 hours from the admission (N=24).

| Variable | Spearman's rho<br>correlation coefficient | P value | Number of<br>observations, N<br>(%) |
| --- | --- | --- | --- |
| IL-6 | 0.42 | 0.0002 | 71 (63.4) |
| D-dimer | 0.62 | 0.0002 | 31 (27.9) |
| LDH | 0.48 | <0.0001 | 82 (73.9) |
| CK | 0.12 | 0.29 | 78 (70.3) |
| Total WBC count | 0.43 | <0.0001 | 87 (78.4) |
| Neutrophil count | 0.50 | <0.0001 | 83 (74.8) |
| Lymphocyte count | -0.1 | 0.38 | 83 (74.8) |
| NLR | 0.44 | <0.0001 | 83 (74.8) |

Legend: IL, interleukin; LDH, lactate dehydrogenase; CK, creatine kinase; WBC, white, blood cell count; NLR, neutrophil to lymphocyte ratio.

Furthermore, the same analysis in the whole cohort by splitting the two group (N=42 for TOCI and N=69 for SOC), showed that baseline CRP value correlated with IL-6, D-dimer, LDH, WBC, neutrophil count and NLR only in the SOC group, while in the TOCI group, baseline CRP correlated only with LDH, WBC, neutrophil and NLR (data not shown).

## Discussion

Our retrospective study was designed to evaluate which baseline standardized laboratory features in hospitalized COVID-19 pneumonia may facilitate optimal employment of experimental anti-cytokine therapy [8, 9]. Some case reports and one case series on the treatment with tocilizumab have been reported in the literature, suggesting some benefits in seriously ill patients [10–16]. More clearly, our data suggested that tocilizumab treatment in patients with cytokine storm features may be more effective outside of the ICU setting in non-ventilated patients. However, there were differences in the degree of inflammation between non-ventilated and ventilated patients treated with tocilizumab, so it cannot be inferred that use of tocilizumab prior to ICU admission is superior, given the generally milder inflammation in the former group. Also, serious superimposed bacterial infections were largely confined to the ICU. More worryingly, half the ICU ventilated patients treated with TOCI remain ventilated or have died with only half of this group showing meaningful clinical improvement so far.

Our findings confirmed that the milder patient group receiving standard of care therapy without the utilization of tocilizumab all made full recoveries. Our findings do point towards trials focused on the earlier use of such therapeutic strategies. Notably, our SOC and TOCI groups were different in terms of co-treatments, which could have affected the overall outcome, and all of the TOCI cases also received antiviral therapy. These findings are preliminary and the results of ongoing randomized controlled trials will definitely clarify anti-cytokine use.

In our study, neutrophilia, lymphopenia, in particular low CD8+ T cell count rather than CD4+ T cell, higher CRP, higher LDH and higher CK showed the highest significance to distinguish the two patient groups at initial hospital admission. Also, serum IL-6 was significantly higher in the TOCI group, thus reflecting the very high inflammatory state of those patients at baseline. Very recently, IL-6 serum levels were also closely correlated with viral load in critically ill patients and it is important to point out that all our patients belonging to TOCI group received anti-viral agents [17]. Notably, baseline CRP and IL-6 continued to distinguish the two groups (TOCI versus SOC) even after excluding the most seriously ill patients from analyses. Thus, these biomarkers could useful for decision making. Notably, a higher NLR, as well as a higher monocyte to lymphocyte ratio, have been associated with mortality and imaging progression in hospitalized patients for COVID-19 [18–20]. It is well known that NLR is a biomarker for poor outcome even in various cancers [21].

Lymphocyte biology probably plays a great role in the pathogenesis of COVID-19 disease [4, 22–25].

Since CD4+ T cells and CD8+ T cells are a crucial arm against infections [26], our findings also indicated that the lymphopenia in the TOCI group may be relevant for secondary infections. Given that, treatment with tocilizumab might favor the persistence of the virus and iatrogenic infections. Anakinra might be safer and more flexible than repeating tocilizumab infusion in seriously ill patients.

A role for anticoagulation is increasingly recognized in severe COVID-19 [27, 28]. In our study, a significant correlation between CRP and D-dimer, as well as with LDH and neutrophil count (and NLR) was shown. Very recent data showed that low molecular weight heparin or unfractionated heparin at prophylactic doses are associated with a reduced short-term mortality in more severe COVID-19 patients [28], and most of our patients, particularly, in ICU, were administered heparin which may have impacted on the overall outcome. Moreover, inflammatory diseases carry a higher risk of thrombosis, as seen in chronic autoimmune diseases [29]. It remains to be seen whether the possible efficacy of anti-cytokine therapy may be even to mitigate against immunothrombosis. Increased levels of LDH and CK may also reflect the level of the organ damage in a systemic disease, as occurs in the macrophage activation syndrome [30], where a hypercoagulable state often complicates the course, and it may be the case for COVID-19. Thus, it is not surprisingly that LDH has been already noticed as biomarker of severity as long as neutrophils, in COVID-19 [3, 31].

This study has several limitations. It is a retrospective study, with some missing data due to the emergency context in which it has been realized. No conclusions on the efficacy and safety of treatment approach employed can be provided. Six patients were transferred from other hospitals so original baseline values from the first admission were unavailable. About 50% of the TOCI group were admitted to the ICU within 24 hours from admission, thus they already presented a more serious disease at the time of admission. The follow-up was limited from our hospital admission to discharge. Finally, measurement of viral load was not available. Nevertheless, the cohort is monocentric and it showed similar characteristics to those reported by Wang et al. [2], thus supporting the results, though preliminary.

To conclude, our study showed that TOCI treated patients COVID-19 pneumonia were at the highest risk of cytokine storm [32]. Tocilizumab use prior to ventilation in ICU may be optimal since 50% of such cases died, remain ventilated and show serious superinfection. Whether the use of tocilizumab prior to ventilation will be vindicated in randomized trials is of major interest. Our findings also showed that cases receiving tocilizumab on ventilation generally had higher levels of inflammation than non-ventilated TOCI treated subjects, possibly suggesting that the latter group has an intrinsically milder disease with a better prognosis. Timing of anti-cytokine therapy is a key issue.

## Data Availability

The data that support the findings of this study are available on request from the corresponding author, [LQ].

## Contributions

LQ designed data collection tools, monitored data collection for the whole study, wrote the statistical analysis plan, cleaned and analysed the data, drafted and revised the paper. He is guarantor. AS collected the data, analysed the data, and revised the paper. FC, MF, TB, ADM, FB, MP, DP collected the data, analysed the data, and revised the paper. SDV, DM analysed the data, drafted and revised the paper. CT designed data collection tools, analysed the data, revised the paper.

## Acknowledgements

We thank the following colleagues for their valued contribution to this work: Ginevra De Marchi, MD, Miriam Isola, prof, BS, Sara Zandonella, MD, Ivan Giovannini, MD, Elena Treppo, MD, Donatella Colatutto, MD, Marco Binutti, MD, Giulia Del Frate, MD, Roberto Agarinis, MD, Valeria Manfrè, MD, Daniela Cesselli, prof, MD, Roberta Giacomello, BS, Federica D’Aurizio, MD, Michele Zuliani, MD, Corrado Marescalco, MD.

## Funding info

This research received no external funding.

## Ethical approval information

The study was conducted in accordance with the ethical principles of the Helsinki Declaration. Patients’ consents for using data for research purpose were obtained at the time of hospital admission.

Ethical approval for the present retrospective observational study was given by “Comitato Etico Unico Regionale (CEUR)”, with the following registration number: CEUR-2020-Os-102. Patients treated with tocilizumab were then enrolled into the observational part of the TOCIVID-19 Italian study (EudraCT: 2020-001110-38), a single arm, open-label trial on the efficacy and safety of tocilizumab in COVID-19 pneumonia.

---

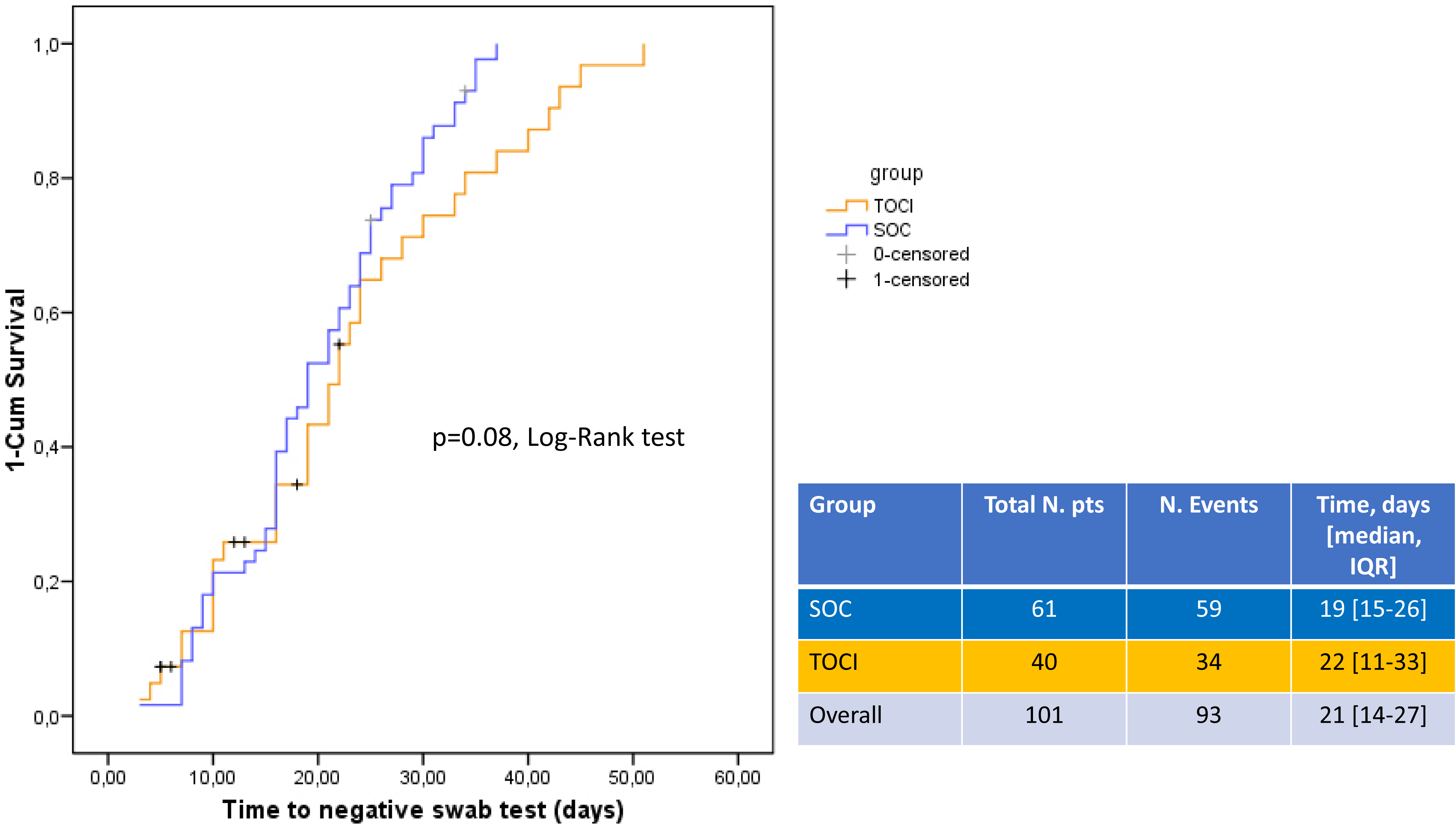

